# Effects of high intensity interval exercise on cerebrovascular function: A systematic review

**DOI:** 10.1101/2020.04.29.20083808

**Authors:** Alicen A Whitaker, Mohammed Alwatban, Andrea Freemyer, Jaime Perales-Puchalt, Sandra A Billinger

## Abstract

High intensity interval exercise (HIIE) improves aerobic fitness with decreased exercise time compared to moderate continuous exercise. A gap in knowledge exists regarding the effects of HIIE on cerebrovascular function such as cerebral blood velocity and autoregulation. The objective of this systematic review was to ascertain the effect of HIIE on cerebrovascular function in healthy individuals. We searched PubMed and the Cumulative Index to Nursing and Allied Health Literature databases with apriori key words. We followed the Preferred Reporting Items for Systematic Reviews. Twenty articles were screened and thirteen articles were excluded due to not meeting the apriori inclusion criteria. Seven articles were reviewed via the modified Sackett’s quality evaluation. Outcomes included middle cerebral artery blood velocity (MCAv) (n=4), dynamic cerebral autoregulation (dCA) (n=2), cerebral de/oxygenated hemoglobin (n=2), cerebrovascular reactivity to carbon dioxide (CO_2_) (n=2) and cerebrovascular conductance/resistance index (n=1). Quality review was moderate with 3/7 to 5/7 quality criteria met. HIIE acutely lowered exercise MCAv compared to moderate intensity. HIIE decreased dCA phase following acute and chronic exercise compared to rest. HIIE acutely increased de/oxygenated hemoglobin compared to rest. HIIE acutely decreased cerebrovascular reactivity to higher CO_2_ compared to rest and moderate intensity. The acute and chronic effects of HIIE on cerebrovascular function vary depending on the outcomes measured. Therefore, future research is needed to confirm the effects of HIIE on cerebrovascular function in healthy individuals and better understand the effects in individuals with chronic conditions. In order to conduct rigorous systematic reviews in the future, we recommend assessing MCAv, dCA and CO_2_ reactivity during and post HIIE.

## Introduction

High intensity interval exercise (HIIE) has emerged at the forefront of exercise regimens due to the shorter activity time needed to benefit[1-3]. HIIE confers similar or significant increased aerobic fitness compared to conventional moderate intensity continuous exercise[1, 4-7]. While aerobic fitness is a measure of increased cardiovascular health, the entire vascular system (including the cerebral vascular system) may be improved following increased aerobic fitness[8]. A review and meta-analysis of HIIE in healthy adults has shown significant increases in aerobic fitness [1, 5, 6, 9]. However, the effects of HIIE on cerebrovascular function have not been systematically reviewed.

Cerebrovascular function is the ability of the cerebral blood vessels to deliver oxygen and nutrients for neuronal metabolism and maintain cerebral blood flow through dynamic autoregulation (dCA). dCA is the ability of the brain to sustain a constant cerebral blood flow despite large fluctuations in peripheral blood pressure[10] [11]. During resting conditions cerebral blood flow responds to arterial blood pressure fluctuations, neuronal metabolism, cortical activation, arterial blood gases and cardiac output [12].

Cerebral blood flow can be measured at rest using magnetic resonance imaging or transcranial Doppler ultrasound (TCD). Middle cerebral artery blood velocity (MCAv) measured by TCD is the only technique to measure cerebral blood flow during exercise, with high temporal resolution [13]. MCAv is linearly related to cerebral blood flow with the caveat that the MCA diameter remains unchanged[14].

A normal cerebrovascular response to submaximal moderate continuous exercise results in increased MCAv [15-17], increased cerebral oxygenation [18, 19] and sustained dCA[20, 21]. MCAv has been shown to concomitantly increase as exercise intensity increases, up to moderate intensity [12, 15, 22-25]. MCAv is affected differently during high intensity exercise. During continuous high intensity exercise and hyperventilation, MCAv is decreased due to a reduction in arterial carbon dioxide (CO_2_)[26, 27] causing downstream arteriole constriction[12, 28]. Cerebrovascular reactivity is the ability of the small vessels in the brain to vasodilate and vasoconstrict in response to fluctuating CO_2_ levels [29, 30]. The cerebrovascular response to HIIE may differ from continuous high intensity exercise due to the repetitive short interval bouts that rapidly increase blood pressure which may cause cerebrovascular hyper-perfusion [31, 32]. If neuroprotective mechanisms of the brain such as dCA do not respond quickly to the repetitive and rapid increases in blood pressure, HIIE could elevate the risk for leakage within the blood brain barrier[31, 33].

Previous scientific statements and narrative reviews have recounted the molecular, hemodynamic and structural processes (i.e. CO_2_, nitric oxide, systemic blood pressure, vessel compliance, glial cell integrity) associated with the cerebrovascular response that may occur during HIIE [33, 34]. However, these detailed narrative reviews[35, 36] did not report the statistical findings of previous studies showing cerebrovascular function during HIIE. To our knowledge, our current systematic review is the first to systematically search and report the results of the dynamic cerebrovascular response during HIIE. Reporting the cerebrovascular response during HIIE is important because it provides objective results to support the previously described narrative statements on hemodynamic processes during HIIE [35, 36]. The purpose of this systematic review was to address the gap in knowledge and report the various study results of HIIE on cerebrovascular function compared to moderate continuous exercise or rest conditions. We systematically examined the results of HIIE studies in healthy individuals based on the operationalization of cerebrovascular outcomes.

## Methods

This review follows the guidelines for Preferred Reporting Items for Systematic Reviews [37]. Literature searches and reviews were performed using PubMed and the Cumulative Index to Nursing and Allied Health Literature (CINAHL) databases. The University of Kansas Medical Center Online Library system was used to access these databases in February, March, and June 2020. In this systematic review, we included peer-reviewed manuscripts written in English from January 2010 to June 2020.

Key words used to search the databases included “high intensity interval training”, “HIIT”, “high intensity interval exercise” “HIIE” AND “cerebral blood flow”, “cerebral blood velocity”, “dynamic autoregulation”. We believe these key words primarily reflect the high intensity interval intervention and cerebrovascular function outcome measures. The main outcomes of this systematic review were MCAv and dCA. However, additional cerebrovascular measures were also included such as oxygenated hemoglobin, cerebrovascular reactivity, cerebrovascular conductance index and cerebrovascular resistance index. Cerebrovascular conductance index is a measure of the conductance of peripheral blood pressure to cerebral blood velocity and is calculated as MCAv/mean arterial pressure (MAP). Cerebrovascular resistance index (MAP/MCAv) measures the resistance of cerebral perfusion pressure to cerebral blood velocity.

The identified abstracts from PubMed and CINAHL were screened using the following inclusion criteria: 1) experimental or quasi-experimental, 2) aerobic exercise identified as the primary means of performing HIIE, 3) cerebrovascular measures were primary or secondary outcomes and 4) human subjects across the lifespan with no current disease. After the removal of duplicates, two researchers screened titles/abstracts for inclusion criteria (A.W. and M.A.). The full texts were examined, and data extracted (A.W. and M.A.). If the authors were unable to come to an agreement, a third author moderated incongruity (A.F.).

A quality review was performed for each article using the modified version of Sackett’s 1981 criteria[38]. We critically analyzed each article’s study design, population, HIIE protocols, cerebrovascular outcomes and results. If an article did not report enough information to determine sufficient quality criteria a “No” rating was given. Articles were rated based on the level of evidence including level I for large randomized control trials, level II for small randomized trial, level III for nonrandomized design, Level IV for case series and Level V for case reports [39].

## Results

The search methods resulted in 67 articles. After removal of duplicates, 15[40-53] articles were identified in PubMed and 5[54-58] new articles in CINAHL. During the initial screening of titles/abstracts, 11 articles were excluded due to HIIE not being the primary experimental protocol performed (n=6), studies not measuring cerebral arteries (n=4) and an animal study (n=1). Studies that combined other interventions with HIIE were excluded due to the confounding variables that could affect cerebrovascular outcomes. After the full text assessment, two articles were excluded due to not meeting experimental or quasi experimental criteria (n=2). See Fig 1 for flow diagram of article selection. [Insert Figure 1]

**Fig 1.**
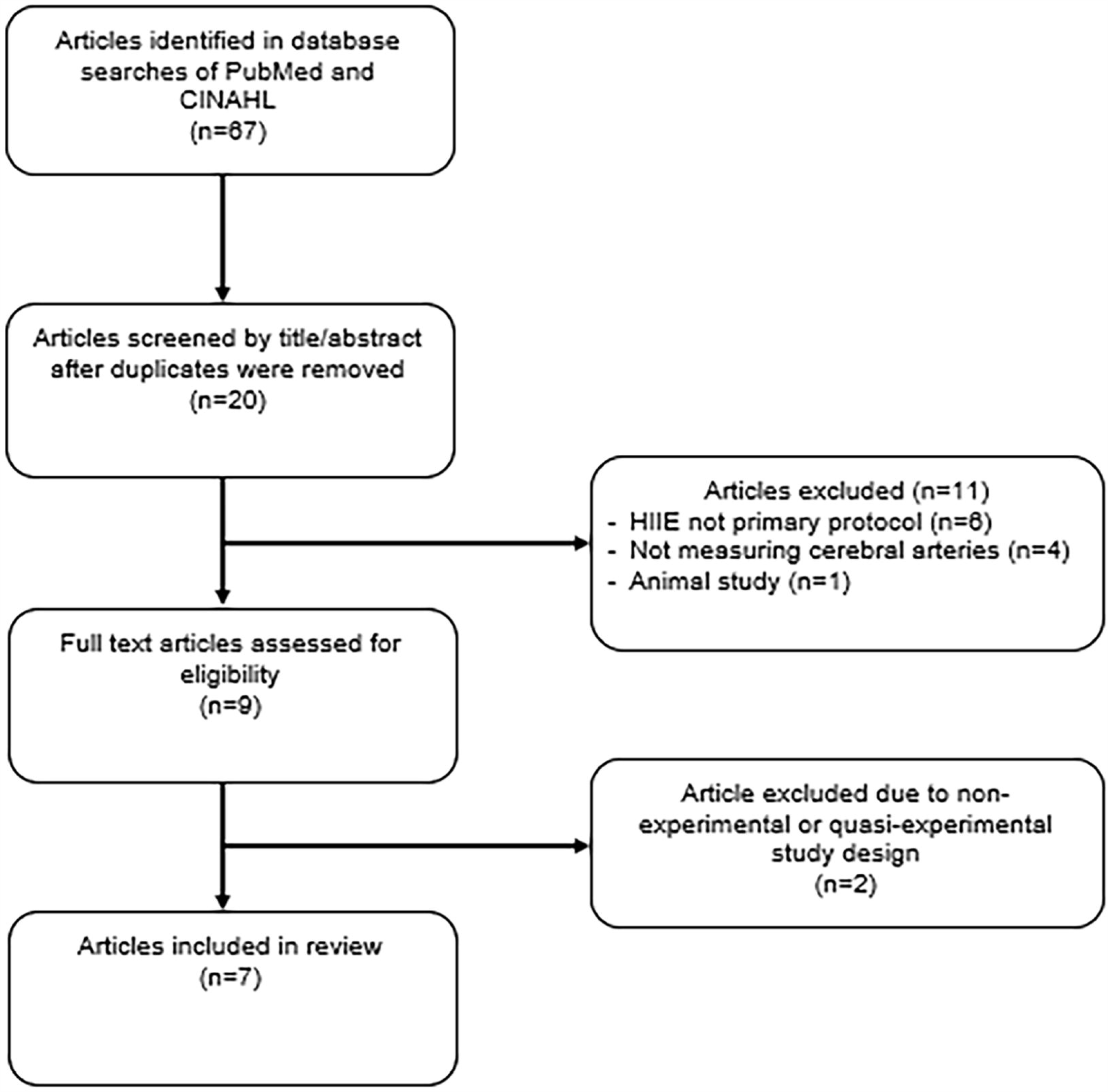
Flow Diagram of Article Selection.

We included seven articles describing cerebrovascular outcomes following HIIE within this review[41, 44, 47, 52, 56, 57, 59]. The full texts are described in Table 1. Of the articles reviewed six were small, randomized trials and one nonrandomized cross-over trial. All the studies involved healthy individuals although some only included men (n=1), women (n=1), or children (n= 1). Prior activity levels of participants ranged from inactive, recreationally active and endurance trained.

**Table 1.** Summary of reviewed articles.

| Study | Design | Level of Evidence | Subjects | Intervention | Outcome measures | Results |
| --- | --- | --- | --- | --- | --- | --- |
| Burma et al, March 2020 | Small Randomized Cross-Over Trial | II | 9 Young Adults | 3 cycling conditions:<br>- High intensity interval training (HIIT, 1-min interval at 85-90% predicted heart rate reserve with 1-min active recovery 15% power output for 10 intervals)<br>- Moderate intensity continuous exercise (MICT, 50-60% predicted heart rate reserve for 45 min)<br>- No-exercise control | Average Exercise MCAv<br>dCA via Transfer Function<br>Analysis during forced MAP oscillations (repeated squat-stand maneuver) | Significant increase in exercise MCAv during MICT compared to HIIT ($p<0.05$) and baseline ($p<0.05$).<br>No change in exercise MCAv during HIIT compared to baseline ($p>0.05$).<br>Significantly higher systolic gain/phase compared to diastolic/mean gain/phase at 0.05 and 0.10 Hz during control ($p<0.05$).<br>Decreased systolic phase in 0.05 Hz immediately following HIIT ($p>0.102$) and MICT until hour 4 |
|  |  |  |  |  |  | (p>0.079).<br>Decreased systolic phase in 0.10 Hz immediately following HIIT until hour 2 (p>.11) and immediately following MICT until hour 4 (p>0.079).<br>No change in gain or coherence in 0.05 Hz or 0.10 Hz during HIIT or MICT. |
| **Burma et al, June 2020** | Secondary Analysis of the above Small Randomized Cross-Over Trial | II | Same as above | Same as above | Cerebrovascular reactivity to hypercapnia<br>Cerebrovascular reactivity to hypocapnia | Significantly decreased absolute and relative MCA reactivity to hypercapnia immediately following HIIT up to hour 2 (p<0.018) compared to MICT and control (p<0.022).<br>Significantly decreased relative MCA reactivity to hypercapnia immediately following MICT up to hour 1 (p<0.024).<br>No significant differences in MCA reactivity to hypocapnia between conditions (p>0.31). |
| **Coetsee et al, 2017** | Small Randomized Controlled Trial | II | 67 Inactive Adults | 16-week intervention Treadmill 30 min, 3x/week<br>4 groups:<br>- High intensity Interval training (HIIT, 4 min interval at 90-95% HRmax with 3 min active recovery 70% HRmax)<br>- Moderate continuous training (MCT, 70-75% HRmax)<br>- No-exercise control (CON). | Cerebrovascular measures taken during cortical activation (Stroop test):<br>- Oxygenated Hemoglobin<br>- Deoxygenated Hemoglobin<br>- Total Hemoglobin Index | No significant differences in oxygenated (effect size=.45, p=.3), deoxygenated (effect size=0.67, p=.14), or total hemoglobin (effect size<.6, p>.18) after HIIT.<br>Significant decrease in deoxygenated hemoglobin (effect size=1.14, p=.01) and total hemoglobin index (effect size=1.49, p<.01) after MCT.<br>Significant increase in oxygenated hemoglobin in CON (effect size=.76, p=.03). |
| **Drapeau et al, 2019** | Small Randomized Clinical Trial | II | 17 Endurance Trained Males | 6-week intervention. Cycled until exhaustion, 3x/week<br>2 groups:<br>- HIIT<sub>85</sub> (1-7 min interval at 85% of maximal aerobic power, with active recovery of 50% of maximal aerobic power)<br>- HIIT<sub>115</sub> (30sec – 1min interval at 115% of maximal aerobic power, with active recovery of 50% of maximal aerobic power). | Resting MCAv<br>Resting CVCi<br>Resting CVRi<br>dCA via Transfer Function Analysis during forced MAP oscillations (repeated squat-stand maneuver) | Significant decrease in phase at 0.10 Hz in HIIT<sub>85</sub> and HIIT<sub>115</sub> (p = .048) with no differences between intensity groups.<br>No significant difference in power spectral density (p > .39), gain (p > .05), or coherence (p>.05) between time or intensity.<br>No significant differences in MCAv (p = .4), CVCi (p = .87), or CVRi (p = .87). |
| **Northey et al, 2019** | Small Randomized Controlled Trial | II | 17 Female Breast Cancer Survivors | 12-week intervention Cycled 20-30 min 3x/week<br>3 groups:<br>- High intensity interval training (HIIT, 30 sec intervals at ~90% maximal heart rate or ~105% peak power with 2 min active recovery)<br>- Moderate intensity continuous exercise (MOD, 55-65% peak power)<br>- No-exercise control (CON) | Resting MCAv,<br>Cerebrovascular Reactivity to CO<sub>2</sub> | No significant differences in resting MCAv (p=.24) or cerebrovascular reactivity (p=.54) after HIIT compared to MOD.<br>No significant differences in resting MCAv (p=.86) or cerebrovascular reactivity (p=.72) after HIIT compared to CON. |
| **Tallon et al, 2019** | Small Randomized Cross-over Trial | II | 8 Prepubertal Children | 2 Cycling conditions:<br>- High intensity interval exercise (HIIE, 1 min interval at 90%max watt with 1 min | Exercise MCAv during each interval<br>Immediate post-exercise MCAv<br>30-minutes post- | Significant decrease in exercise MCAv during the 6<sup>th</sup> interval of HIIE compared to baseline (10.7%, p=.004).<br>Significant decrease in exercise MCAv during |
|  |  |  |  | active recovery at 20%max watt for 6 intervals)<br>- Moderate-intensity steady-state exercise (MISS, 15 min at 44%max watt) | exercise MCAv | the 3<sup>rd</sup> and 4<sup>th</sup> intervals of HIIE compared to MISS (p=.001). Significant decrease in MCAv immediately post-exercise following HIIE and MISS (p<.001).<br>No significant difference in MCAv at 30-minutes post-exercise following HIIE and MISS compared to baseline (p>.05).<br>Significant increase in exercise MCAv during the 2<sup>nd</sup> minute of MISS compared to baseline (5.8%, p=.004). |
| **Monroe et al, 2015** | Nonrandomized Cross-Over Trial | III | 15 Recreationa lly Active Adults | 2 cycling conditions:<br>- Sprint Interval Cycling (SIC, 30 second all-out sprint interval with 4 min active recovery for 4 intervals)<br>- Constant Resistance Cycling (CRC, 18 min at 70rpm with resistance set by matching total work performed during SIC) | Oxygenated Hemoglobin (HbO2)<br>Deoxygenated Hemoglobin (HHb) | Significant increase in average HbO2(effect size=.536, p=.001), minimum HbO2 during recovery (effect size=.392, p<.001) and maximum HbO2 during recovery (effect size=.588, p=.001) in SIC compared to CRC.<br>Significant increase in average HHb during SIC compared to CRC (effect size=.386, p=.003). |

MCAv = middle cerebral artery blood velocity, dCA = dynamic cerebral autoregulation, min = minute,
HRmax = maximum heart rate, CVCi = cerebrovascular conductance index, CVRi = cerebrovascular
resistance index, CO<sub>2</sub> = carbon dioxide

### High intensity interval protocols

Methods of prescribing HIIE varied greatly between studies. HIIE protocols included 6-to 16-week exercise interventions (n = 3) or one single bout of exercise(n=4). By examining 6-to 16-weeks of HIIE, the long-term or chronic effects of this intervention were studied. By examining a single bout of HIIE, the immediate or acute effects of the exercise were reported. In addition to the duration variability, we found that the mode of HIIE also differed across the included studies. One study used a treadmill as the mode of exercise with 4-minute intervals of 90-95% maximal heart rate for 30 minutes. The remaining six studies used cycling as the mode of exercise but differed in parameters ranging from 30 seconds to 7-minute intervals at 85% to 115% of maximal watts or ∼ 85% to 90% maximal heart rate. A constant between all studies included an active (rather than passive) recovery interval between sprints. However, the intensity and duration of recovery intervals differed greatly.

### Cerebrovascular outcome measures

The results of this review can be operationalized based on the outcome variables measured during HIIE such as MCAv (n=4), dCA (n=2), cerebral de/oxygenated hemoglobin (n=2), cerebrovascular reactivity to CO_2_ (n=2) and cerebrovascular conductance/resistance index (n=1). Table 2 describes whether HIIE increased, decreased or had no influence on the operationalized cerebrovascular measures. A meta-analysis was not performed due to low number of studies (≤ 2) reporting each operationalized cerebrovascular measure.

**Table 2.**
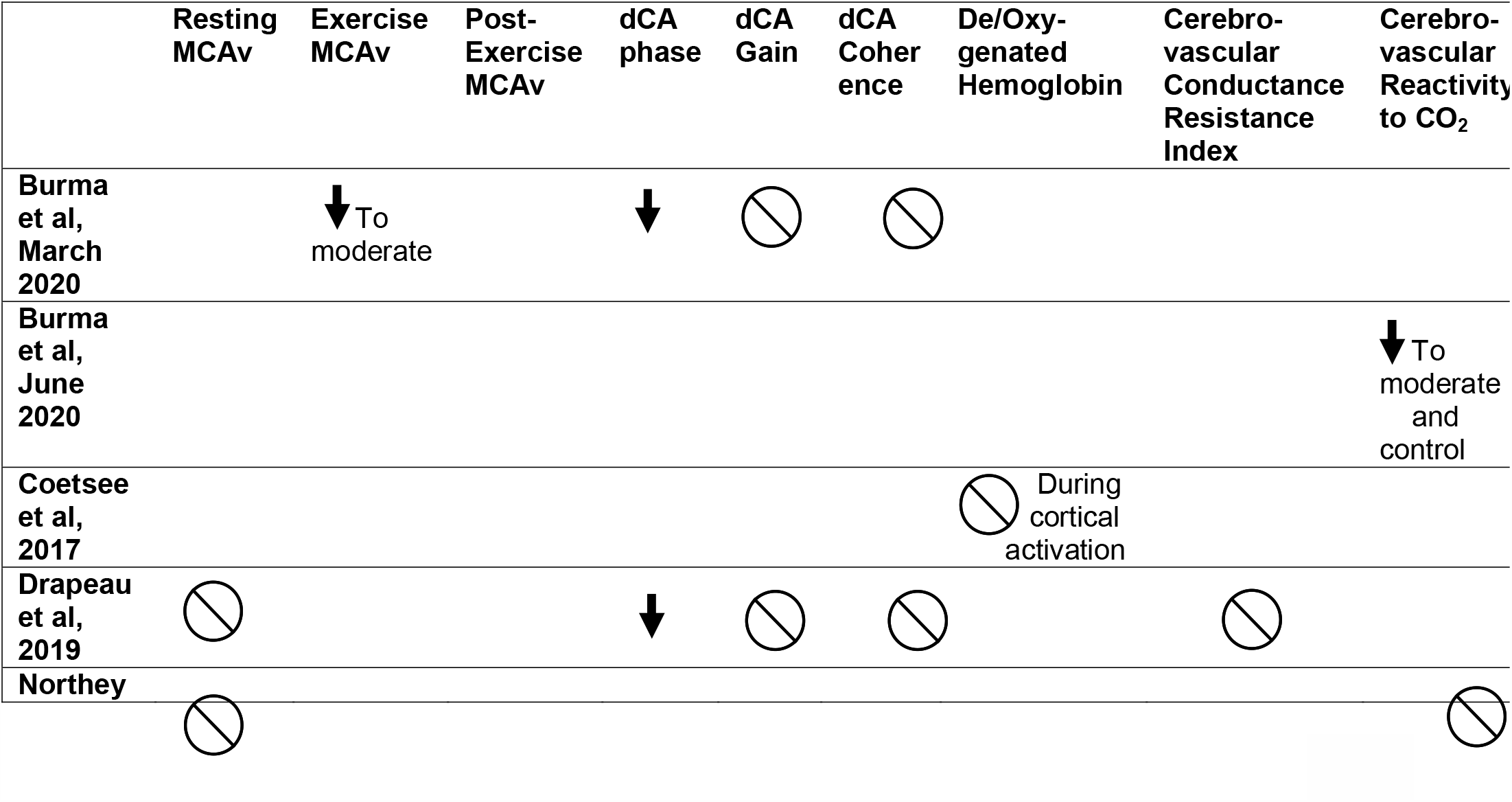
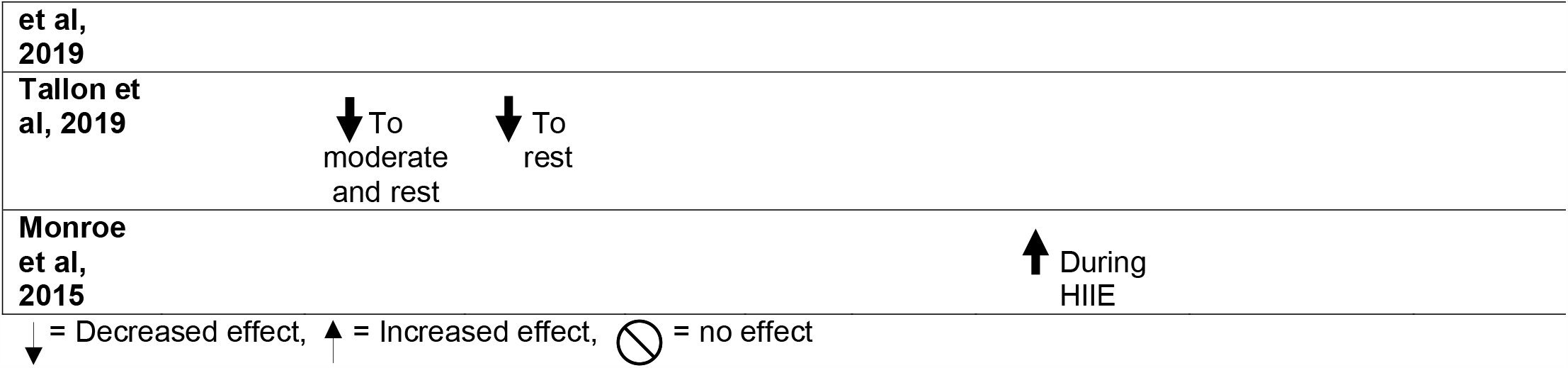
Summary of the effects of HIIE on operationalized cerebrovascular measures.

#### MCAv

Of the studies reporting MCAv outcomes, resting MCAv (n = 2), exercise MCAv (n = 2) and MCAv immediately post exercise (n = 1) were used. No significant differences were found for resting MCAv after 6- or 12-weeks of HIIE when compared to moderate continuous exercise or control[41, 44]. During HIIE, exercise MCAv was significantly decreased compared to moderate continuous exercise[56, 59].

Conflicting results were found between two studies comparing exercise MCAv to rest. Burma et al.[59] reported no significant difference between average exercise MCAv and rest in adults. However, rather than reporting average exercise MCAv of the entire HIIE bout, Tallon et al.[56] reported exercise MCAv for each 1-minute sprint interval of HIIE. During the 6th sprint interval of HIIE, Tallon et al.[56] reported significantly decreased exercise MCAv compared to rest which remained immediately following exercise[56].

#### dCA

Transfer function analysis of dCA was reported in the very low and low frequency bands (n = 2). Drapeau et al.[41] conducted a 6-week intervention of HIIE and reported a significant decrease in phase compared to rest with no significant change in coherence or gain. Burma et al.[59] conducted a single bout of HIIE and reported decreased MCAv systolic phase immediately following exercise that extended up to four hours later.

#### De/oxygenated hemoglobin

Oxygenated and deoxygenated hemoglobin were reported during a single bout of HIIE (n=1) and during a 16-week HIIE intervention (n=1). Monroe et al.[57] conducted a single bout of HIIE and reported an increase in oxygenated and deoxygenated hemoglobin during HIIE compared to moderate continuous exercise. Coetsee et al.[47] conducted a 16-week intervention of HIIE and reported no significant lasting changes in oxygenated or deoxygenated hemoglobin during cortical activation.

#### Cerebrovascular reactivity

Cerebrovascular reactivity to CO_2_ were reported during a single bout of HIIE (n=1)[52] and during a 12-week HIIE intervention (n=1)[44]. After a single bout of HIIE, cerebrovascular reactivity to higher CO_2,_ or hypercapnia, was significantly decreased by 37% and remained an hour later. The reduced cerebrovascular reactivity to hypercapnia was also significantly different than moderate intensity and control. Cerebrovascular reactivity to lower CO_2_, or hypocapnia, was not significantly different following a single bout of HIIE. Cerebrovascular reactivity to CO_2_ was also not significantly different following 12 weeks of HIIE[44].

#### Cerebrovascular conductance and resistance

Cerebrovascular conductance index and cerebrovascular resistance index were only reported in a single study each. A 6-week HIIE intervention reported no significant changes in cerebrovascular conductance index or cerebrovascular resistance index[41].

### Quality review

The quality review of each study is presented in Table 3. Out of seven total quality criteria, (n=2) reported five quality criteria, (n=1) reported four quality criteria and the remaining (n=4) reported three quality criteria. Therefore, the overall quality criteria results were moderately poor. All studies accounted for subjects and monitored the HIIE protocol parameters. No studies reported avoidance of contamination or co-intervention. No studies reported blinding of the outcome assessments. Only Burma et al.[52, 59] and Monroe et al.[57] reported their reliability via coefficient of reproducibility and intraclass coefficients of their measures. And only Burma et al.[52, 59] and Drapeau et al.[41] reported validity of their respective cerebrovascular outcomes.

**Table 3.** Summary of quality review.

|  | **Avoided Contamination and Co-Intervention** | **Random Assignment to Conditions** | **Blinded Assessment** | **Monitored Intervention** | **Accounted for All Subjects** | **Reported Reliability of Measures Used** | **Reported Validity of Measures Used** | **Total Number of Criteria Met** |
| --- | --- | --- | --- | --- | --- | --- | --- | --- |
| **Burma et al, March 2020** | No | Yes | No | Yes | Yes | Yes | Yes | 5 |
| **Burma et al, June 2020** | No | Yes | No | Yes | Yes | Yes | Yes | 5 |
| **Coetsee et al, 2017** | No | Yes | No | Yes | Yes | No | No | 3 |
| **Drapeau et al, 2019** | No | Yes | No | Yes | Yes | No | Yes | 4 |
| **Northey et al, 2019** | No | Yes | No | Yes | Yes | No | No | 3 |
| **Tallon et al, 2019** | No | Yes | No | Yes | Yes | No | No | 3 |
| **Monroe et al, 2015** | No | No | No | Yes | Yes | Yes | No | 3 |

## Discussion

This review met the objective of reporting the results of various HIIE studies and the effects on operationalized cerebrovascular function in healthy individuals. This review is the first to report the effects HIIE on cerebrovascular function compared to moderate continuous exercise and rest in healthy individuals. In general, we found that the acute and chronic effects of HIIE on cerebrovascular function vary largely depending on the methods and outcomes measured.

### MCAv

In these studies, 6-to 12-week HIIE interventions had no effect on resting MCAv in healthy individuals. No significant change may be from too short of a HIIE intervention duration. Also a ceiling effect may be observed for young, healthy individuals and could explain no changes in resting MCAv[16]. During a single bout of HIIE, hyperventilation and downstream arteriole vasoconstriction may explain the acute decreases in exercise MCAv compared to moderate continuous exercise[12, 28, 60, 61]. Vasoconstriction may play a protective role during HIIE due to heightened peripheral blood pressure potentially causing hyper-perfusion or damage to the blood brain barrier[10, 33, 62]. During a single bout of HIIE, there is contradictory evidence comparing exercise MCAv to rest. One study reported no change in average exercise MCAv compared to resting. Another study reported decreased exercise MCAv after six sprint intervals of HIIE and remained decreased compared to rest immediately following HIIE[56]. The differences reported in exercise MCAv compared to rest could be due to age (adults versus prepubertal children) or due to the analysis of MCAv during HIIE (average over entire exercise versus separate sprint intervals). Decreases in exercise MCAv compared to rest may only occur in the late intervals of HIIE, during hyperventilation. Therefore, exercise MCAv should be reported for each interval of HIIE rather than an average of the entire exercise bout.

### dCA

After a 6-week intervention and single bout of HIIE, dCA phase was decreased compared to rest. The chronic effects of HIIE on dCA phase may be due to elevated cardiorespiratory fitness in endurance trained individuals being associated with attenuated dCA [41, 63]. In healthy individuals, increased frequency within MCAv and MAP waveforms (that can occur with HIIE) may cause a reduction in phase due to dCA being a high-pass filter[64, 65]. Burma et al.[59] also suggests that systolic phase may reveal greater changes in dCA than both diastolic and mean phase. After a single bout of HIIE, reduction in systolic phase extended up to 4 hours and therefore the common approach of abstaining from exercise 12 hours before research studies[66-68] may be too conservative[59].

Although not included in this review due to the observational study design, contradictory evidence of sustained dCA during HIIE has been reported[45]. Differences in exercise parameters between HIIE may be the cause to contradictory findings due to exhaustive exercise showing decreased dCA [31, 69]. More studies are needed to confirm the acute and chronic decreases in dCA following HIIE.

#### De/oxygenated hemoglobin

After a 16-week HIIE intervention, oxygenated and deoxygenated hemoglobin during cortical activation did not change. However, the 16-week HIIE intervention decreased reaction time during cortical activation and therefore may have increased efficiency of cortical oxygen use. Also, during a single bout of HIIE oxygenated and deoxygenated hemoglobin increased compared to moderate continuous exercise. The acute and chronic effects of HIIE on oxygenated and deoxygenated hemoglobin still needs further investigation due to each only being reported in a single study.

#### Cerebrovascular reactivity

A 12-week HIIE intervention did not significantly change cerebrovascular reactivity which could be due to vascular desensitization from chronic exposure to CO_2_ during HIIE[33, 70]. Following a single bout of HIIE, cerebrovascular reactivity to hypercapnia was decreased showing the inability of the cerebrovascular system to maximally vasodilate. The maximal capacity for vasodilation after HIIE may be reduced following HIIE due to prolonged cerebrovascular vasoconstriction that occurs with hyperventilation during HIIE[33, 52]. Cerebrovascular reactivity to hypocapnia was not changed following a single bout of HIIE due to the ability of the vessels to vasoconstrict remaining intact[52]. The reduction in cerebrovascular reactivity to higher CO_2_ remains an hour after HIIE. Therefore, the authors conclude again that the common approach of abstaining from exercise 12 hours before research studies[67, 71, 72] may be too conservative[52].

#### Cerebrovascular conductance and resistance

Cerebrovascular conductance and resistance were not significantly changed following a 6-week HIIE intervention. While a 6-week HIIE intervention significantly improved peripheral arterial conductance and resistance[4], this change in the peripheral arteries may not be demonstrated in the cerebrovascular arterial conductance or resistance[35, 41]. However, due to the cerebrovascular conductance or resistance index being reported in only a single study, no conclusive effects of HIIE can be determined.

### Article limitations

The individual study limitations have been previously acknowledged within the published articles or have been identified by authors (AW and MA) during the quality review. Most of the studies were small, randomized trials and included mostly young individuals. The small sample size available in males (n = 17), females (n = 17), or children (n = 8) limit the generalizability of results presented in this systematic review to a larger population. All studies implemented HIIE protocols that differed in time, repetitions and intensity levels, therefore making comparisons between studies difficult. These studies report potential limitations to the methods of measuring cerebrovascular function. TCD is the preeminent method of measuring MCAv during exercise but MCAv during HIIE may be underestimated[73]. However, some of the studies report the minor changes in expired CO_2_ during HIIE were not likely to induce changes in MCA diameter[14, 56, 57]. The studies also report underestimation of cerebral oxygenation due to the two-channel near infrared spectrometer not measuring the motor, occipital, or parietal cortex [47, 57].

### Review limitations

The authors acknowledge a risk of publication bias by only including peer-reviewed articles written in English and did not include grey literature. The cerebrovascular function measures included within this review vary greatly and have vast heterogeneity. While HIIE is not a new mode of exercise, studying cerebrovascular measures during HIIE is novel. Therefore, authors could only identify seven small studies with the oldest article dating back to 2015. The primary outcome of MCAv (n=4) and dCA (n=2) were reported in few studies with low power. Therefore, a meta-analysis could not be performed due to insufficient mathematical combination.

## Conclusion

This review has provided preliminary information studying the effects of HIIE on cerebrovascular function. Although the amount of studies reporting cerebrovascular function during HIIE was small, we propose that a single bout of HIIE may acutely decrease MCAv, dCA and cerebrovascular reactivity to hypercapnia while increasing oxygenated hemoglobin, during or immediately following HIIE compared to moderate continuous exercise or rest conditions. No significant chronic effects were found for resting MCAv, cerebrovascular reactivity to CO_2_, cerebrovascular conductance or resistance index after 6- or 12-week interventions of HIIE. Further research is warranted to confirm the acute and chronic effects of HIIE on MCAv, dCA, and cerebrovascular reactivity.

With increased interest in healthy brain aging and implementing interventions to maintain or improve brain health[74], studying the effects of HIIE interventions are critically needed[35, 36]. While this review only included healthy individuals, we provide an early reference to understanding “normal” physiological effects of HIIE on cerebrovascular function and the need to compare to clinical populations.

Future HIIE research is needed to evaluate cerebrovascular function not only during HIIE but also immediately following exercise. More studies such as randomized controlled trials with large sample sizes are needed to conduct a meta-analysis to combine and statistically analyze the summary results of HIIE on cerebrovascular function. Additionally, more studies are needed to determine the optimal interval parameters for HIIE and cerebrovascular function for humans.

## Data Availability

Available data is included within the systematic review and within the previously published original manuscripts.

## Supporting Information

S1 Fig. Prisma flow diagram

S1 Table. Prisma checklist

